# Neutrophil Lymphocyte Ratio as a Predictor of Glucocorticoid Effectiveness in Covid-19 Treatment

**DOI:** 10.1101/2021.06.15.21251794

**Authors:** Benjamin J. Lengerich, Rich Caruana, Alex Peysakhovich, Leora Horwitz, Yin Aphinayanaphongs

## Abstract

Glucocorticoids have been shown to improve outcomes of patients with severe cases of Covid-19. However, criteria for prescribing glucocorticoids are currently limited. To identify potential for targeting, we perform an observational analysis of mortality of hospitalized patients. Our results agree with current clinical understanding that glucocorticoids benefit patients with severe cases of Covid-19, and that elevated Neutrophil/Lymphocyte Ratio (NLR) is associated with mortality. Furthermore, our results suggest that glucocorticoids could be targeted to patients with elevated NLR (especially in the range 6-25) at time of admission. Finally, we note there are also high-risk patients with low NLR, suggesting varying presentations of severe Covid-19.

---

Glucocorticoids have been shown to improve outcomes of patients with severe cases of Covid-19 [1]. However, criteria for prescribing glucocorticoids are currently limited. To identify potential for targeting, we perform an observational analysis of patient mortality. Our results suggest that glucocorticoids could be targeted to patients with elevated Neutrophil/Lymphocyte Ratio (NLR) at time of admission.

Our outcome is in-hospital mortality. We define “glucocorticoid treatment” as treatment with a glucocorticoid within 24 hours of hospital admission; this captures 193 of 237 (81.4%) patients who took a glucocorticoid at any point in their hospitalization. Our control condition is not receiving any glucocorticoids in the first 24 hours. Our analysis includes deidentified records from 3108 patients hospitalized for Covid-19 with 193 receiving glucocorticoids. This cohort includes patients hospitalized from March to August 2020 with an average mortality rate of 18.1%. Mortality rate peaked over 25% and decreased to less than 5% in August.

Treatment assignment is not random and has changed over time with more glucocorticoid prescriptions at later dates [2]. To correct for this and other confounding, we use a two-stage machine learning procedure to estimate the adjusted risk ratio (ARR) [3, 4], which can be interpreted as an odds ratio after correcting for patient mortality risk at admission. Our two-stage procedure works as follows: first we take half of the sample and train a generalized additive model (GAM) [5, 6] to predict mortality from patient features at admission. In our held-out sample, we compute the excess risk after accounting for the model predictions of patient risk. To compute the effect of glucocorticoids, we compare the average excess risk in glucocorticoid patients with the average excess risk in the control patients. To compute standard errors and p-values, we bootstrap the held-out sample. The baseline mortality risk model achieves AUC 0.91 on held-out patients. See S3 for details of model construction.

After correcting for confounding variables, we estimate no average treatment effect (ARR= 0.96, 95% confidence interval (CI) 0.86-1.09 (S4)). However, additional analysis finds that glucocorticoids have heterogeneous effects, i.e. glucocorticoids do not benefit all patients equally.

The importance of NLR in the baseline mortality risk model (S3) and previous analyses of severe Covid-19 [7] led us to stratify our analysis by patient NLR. Because glucocorticoids can affect patient NLR, we use only initial lab tests which are taken before in-patient medications are administered. We estimate ARRs for three ranges of NLR: 0-6, 6-25, and >25. We chose these ranges by looking at the predictions of risk from our GAM on the training set (Figure 1) and breaking the continuous NLR variable into 3 regimes.

**Figure 1:**
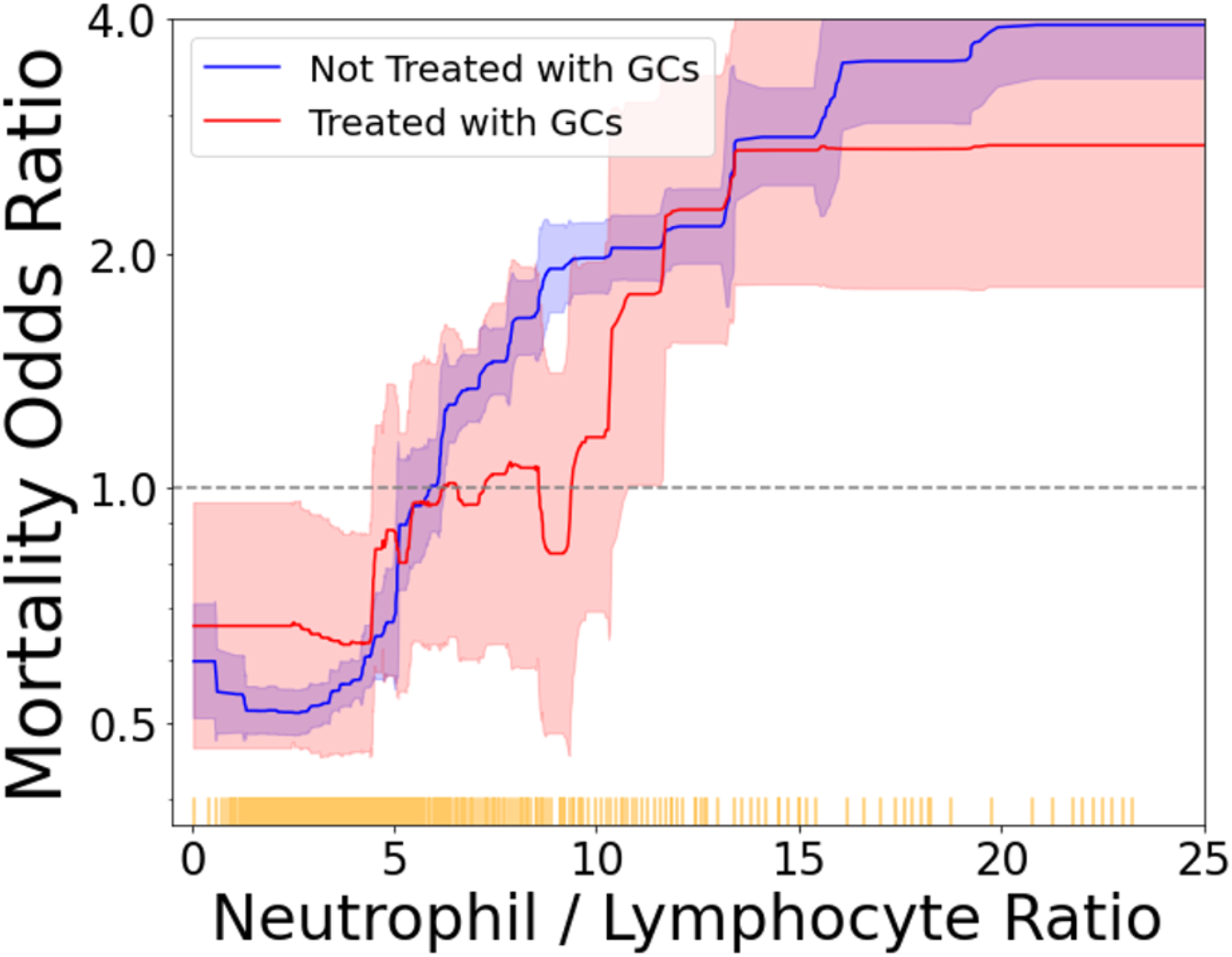
Mortality risk conferred by NLR in addition to all other risk factors, with patients stratified by glucocorticoid treatment. We plot the risk curve estimated by the generalized additive model, with 95% confidence intervals shaded. Each yellow tick mark along the horizontal axis indicates 10 patients.

Corresponding sample sizes and ARRs are given in Table 1. Of these three ARRs, we observe statistically significant evidence of glucocorticoid effects only for patients with NLR 6-25. We hypothesize that for patients who are not at risk of severe inflammation, glucocorticoids have little effect; for patients who are admitted with extremely high NLR, glucocorticoids may be insufficient. Though ARR is standard, we also calculate adjusted risk differences (S4) for another view of the benefit of glucocorticoids.

**Table 1:** Adjusted Risk Ratios (ARRs) of mortality for patients treated with glucocorticoids compared to patients not treated with glucocorticoids, calculated by patient NLR. ARRs smaller than 1 indicate reduced mortality for patients treated with glucocorticoids, i.e., a beneficial effect. P-values are calculated by bootstrap resampling of the test set.

| NLR Values | N GC | N Control | ARR | 95% CI | P-Value |
| --- | --- | --- | --- | --- | --- |
| 0-6 | 79 | 1728 | 0.99 | 0.68-1.31 | 0.975 |
| 6-25 | 99 | 1079 | 0.74 | 0.49-0.99 | **0.042** |
| >25 | 15 | 116 | 0.90 | 0.58-1.22 | 0.549 |

The results agree with current clinical understanding that glucocorticoids benefit patients with severe cases of Covid-19, and that elevated NLR is associated with mortality (S3). However, there are also high-risk patients with low NLR (S3), suggesting varying presentations of severe Covid-19, some of which may not respond to glucocorticoids. Our study comes with all caveats of observational analyses, and we suggest randomized control trials to study heterogeneous treatment protocols.

## Data Availability

Data include anonymized medical records which are not publicly available.

