## Supplement for "Neutrophil Lymphocyte Ratio as a Predictor of Glucocorticoid Effectiveness in Covid-19 Treatment"

#### S1 Dataset Construction

Our dataset consists of 11080 total hospitalized patients who have lab-confirmed cases of Covid-19. To filter out patients who were hospitalized for reasons other than Covid-19, we excluded patients who have indicators of (1) pregnancy: outpatient prenatal vitamins, in-patient oxytocics, folic acid preparations; or (2) scheduled surgery: urinary tract radiopaque diagnostics, laxatives, general anesthetics, antiemetic/antivertigo agents, or antiparasitics. We also require that the patients have recorded temperature, age, BMI, and Admission Day. Finally, we remove patients who died within six hours of admission.

To correct for patient risk confounding, we observe pre-admission features including demographics, comorbidities, outpatient medications, initial in-patient vitals, and initial in-patient lab tests. We exclude any measurement taken within 24 hours of the patient mortality. The 45 total features are listed below.

- Demographics/Vitals:
  - Age
  - Sex
  - BMI
  - Day
  - Temperature
- Comorbidities:
  - Myocardial Infarction
  - Congestive Heart Failure
  - Peripheral Vascular Disease
  - Cerebrovascular Disease
  - Dementia
  - Chronic Obstructive Pulmonary Disease
  - Peptic Ulcer Disease
  - Mild Liver Disease
  - Diabetes without chronic complications
  - Diabetes with chronic complications
  - Hemiplegia or paraplegia
  - Renal disease
  - Cancer (any malignancy)
  - Metastatic solid tumor
  - Charlson score
  - Hypertension
  - Atrial fibrillation
  - Valve Replacement
  - Rheumatoid Arthritis
- Outpatient Medications taken before hospitalization (limited to medication classes taken by at least 100 patients):
  - Antihyperglycemic, Biguanide Type
  - Laxatives And Cathartics

  - Platelet Aggregation Inhibitors
  - Vitamin D Preparations
  - Calcium Channel Blocking Agents
  - Proton-Pump Inhibitors
  - Antihyperlipidemic-Hmgcoa Reductase Inhib (Statins)
  - Beta-Adrenergic Agents, Inhaled, Short Acting
  - Blood Sugar Diagnostics
  - Anticonvulsants
  - Analgesic/Antipyretics,Non-Salicylate
  - Beta-Adrenergic Blocking Agents

- Lab Values:
  - Potassium (0.5% Missing)
  - Ferritin (8.3% Missing)
  - Calcium (0.5% Missing)
  - Neutrophil % (0.0% Missing)
  - Lymphocyte % (0.0% Missing)

Neutrophil-Lymphocyte Ratio is calculated by dividing Neutrophil % by Lymphocyte %. We exclude all patients who are missing either of these lab values.

The patient population has changed over time. The majority of patients were admitted from days 30-50. As Figure S1 shows, this period also contained the majority of patients with extremely elevated NLR levels. However, the range of NLR observed in the days and months following the initial peak of the pandemic remains wide. Many factors have shifted over the course of the pandemic, so we also control for patient intake day in our models.

### S2 Treatment and Outcome Measure Construction

In this study, we are interested in the use of glucocorticoids. To ensure a proper linking of lab values, treatments, and outcomes, we consider only glucocorticoid treatments that are given within 24 hours of the initial lab test and at least 24 hours before mortality. In this set of patients, the number of doses by glucocorticoid medication are: 627 Methylprednisolone, 291 Prednisone, 186 Hydrocortisone, 165 Dexamethasone. Figure S2 shows that the proportion of Covid-19 patients being prescribed glucocorticoids has increased over time from a minimum of less than 5% to a more recent peak near 30%.

Glucocorticoid prescriptions are correlated with: Admission Day ($R = 0.16$), Chronic Obstructive Pulmonary Disease ($R = 0.13$), Outpatient Beta-Adrenergic Agents ($R = 0.11$), Valve Replacement ($R = 0.10$), and Increased Charlson Score ($R = 0.10$). Glucocorticoid prescriptions are anti-correlated with: In-Patient Hydroxychloroquine ($R = -0.06$), Hematocrit ($R = -0.05$), and In-Patient Analgesic/Antipyretics ($R = -0.05$).

### S3 Mortality Risk Model

We use generalized additive models (GAMs) to model patient mortality risk at admission. GAMs are a version of logistic regression that are able to model non-linear effects [5]. While logistic regression summarizes the influence of each feature with a single coefficient, GAMs estimate the influence of a feature for every value the feature can take on as a graph. This means that GAMs naturally accommodate non-linear effects, which improves both model accuracy and interpretation. Modeling non-linear effects is particularly important when features have multiple regions of high or low risk (e.g., both hyperthermia and hypothermia are associated with high risk).

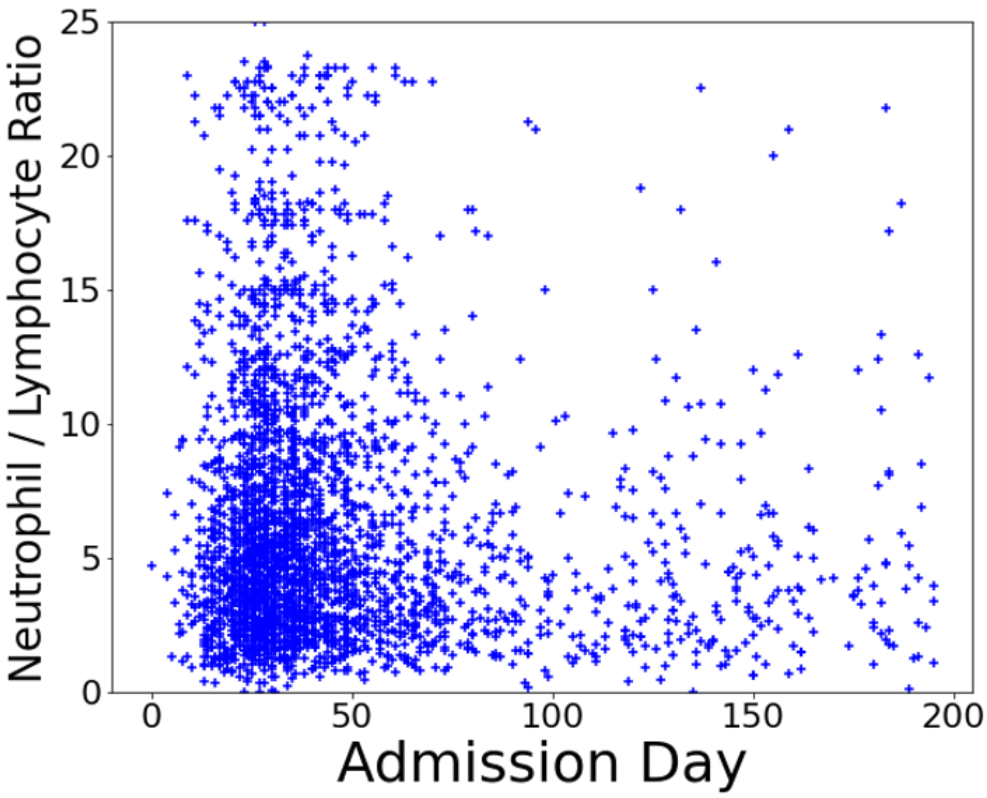


Figure S1: Distribution of patients by NLR and Admission Day. Each marker indicates a single patient, with the vertical axis indicating NLR value on the initial lab test and the horizontal axis indicating the day the patient was admitted.

We use tree-based GAMs [6] implemented in the Python `Interpret` package[1]. These GAMs are invariant to all monotonic feature transforms, so log-transforms of lab values are not necessary.

The risk model achieves an ROC of $0.912 \pm 0.001$ and an F1-score of $0.598 \pm 0.002$ on held-out patients. This significantly outperforms a logistic regression model which achieves an ROC of $0.859 \pm 0.001$ and F1-score of $0.455 \pm 0.002$ on the same data.

The 5 most important features to this background risk model are:

1. Temperature
2. Age
3. Admission Day
4. Calcium
5. Charlson score

If NLR is included in this model, it jumps to becoming the most important feature. Figure S4 demonstrates the effect of NLR on mortality risk estimated in this model.

While elevated NLR is a strong predictor of mortality, it is not the only risk factor. We can summarize all of these risk factors through the GAM risk model. Figure S4 shows that there are many patients with low NLR levels who nevertheless have large probabilities of mortality, and many of these patients died.

[1] https://github.com/interpretml/interpret

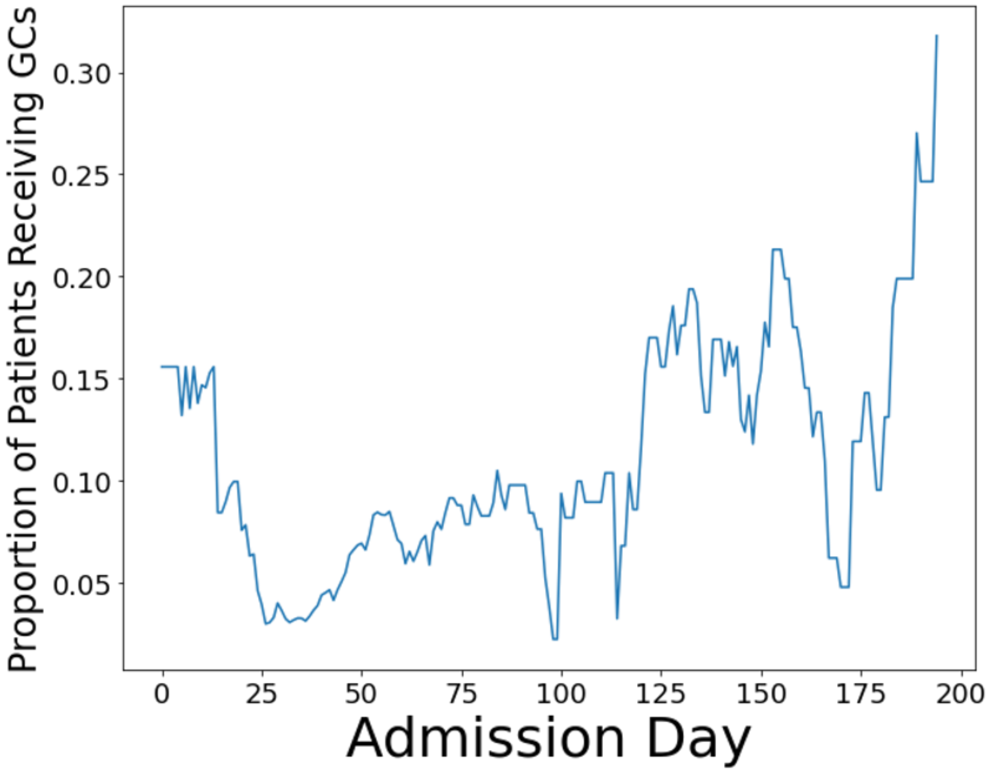


Figure S2: The proportion of patients being prescribed glucocorticoids (GCs) has increased over time. The values are smoothed over a 14-day period by a running average.

#### S4 Interpretation of Main Results

Glucocorticoid prescription is highly correlated with later admission dates and mortality is lower at later dates for a number of reasons. To correct for this confounding, as well as confounding from patient characteristics, we use the mortality risk model described in S3 to correct for all risk which can be attributed to factors other than glucocorticoids.

We are interested in

$$P_1(x, y) = P(\text{mortality}|X = x, \text{NLR} = y, \text{Glucocorticoid} = 1),$$
$$P_0(x, y) = P(\text{mortality}|X = x, \text{NLR} = y, \text{Glucocorticoid} = 0)$$

. Comparing $P_1$ and $P_0$ separates mortality risk that can only be assigned to glucocorticoid treatment out from mortality risk which could be assigned to other risk factors.

We follow the convention of [4] to estimate these quantities using a classification model. We use the additive model assumption that $P(\text{mortality}|X = x, \text{NLR} = y, \text{Glucocorticoid} = z) = f(x) + g(y, z)$. This means that a single $f$ (the background risk GAM trained in S3) is shared between both groups of patients and that $g$ is an adjustment to this background risk model.

For a small set of NLR values, we can estimate each $g(y_i, z_j)$ as the residual of the predictions of the background risk model on a set of held-out patients with NLR= $y_i$ and Glucocorticoid= $z_j$. However, for the continuous-valued NLR, we need a model to provide a smooth estimate of g. For this, we use a tree-based interaction available in `Interpret`. This whole procedure can be run as a native operation for the EBM model in `Interpret`, which trains and freezes main effects before estimating interaction effects. For robustness, we estimate $g$ on a test set of held-out patients which is resampled in each bootstrap iteration.

The adjusted risk ratio (ARR) is the ratio $\frac{P_1}{P_0}$, while the adjusted risk difference (ARD) is the difference $P_1 - P_0$

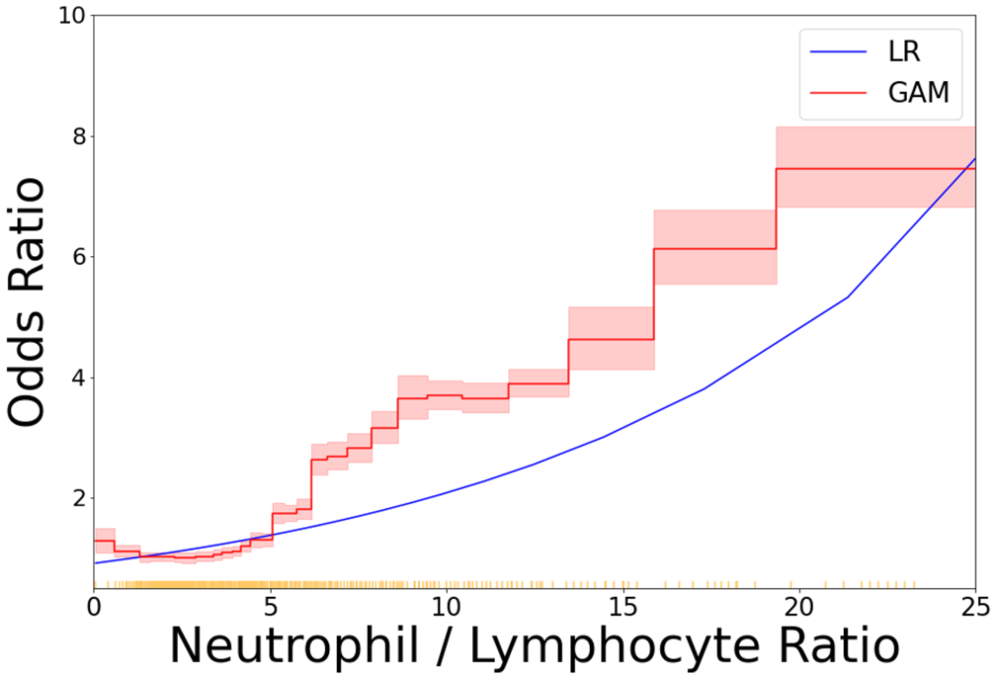


Figure S3: Mortality risk conferred by NLR in addition to all other risk factors, for patients not treated with glucocorticoids. We plot the risk curve estimated by the generalized additive model (red), and a logistic regression model (blue). Each yellow tick mark along the horizontal axis indicates 10 patients. Unlike the logistic regression model, the GAM suggests risk rises rapidly beginning at NLR=6.

[3, 4]. For both ARR and ARD, lower values indicate protective effects of glucocorticoids. The ARR provides the protective effect as a relative measure while the ARD provides the absolute difference in risk between the groups.

We first use this procedure to estimate the effect of glucocorticoids as a homogeneous benefit to all Covid-19 patients. We do not observe a strong benefit: mean ARR 0.96 (95% CI 0.86-1.09). Thus, we examine possible heterogeneous effects of GCs.

We examine the heterogeneous effect of glucocorticoids with benefit modulated by NLR level. Due to the limited sample size for patients with NLR>25, we focus our analysis on patients with NLR<25. Figures S5 and S6 show that the benefit of glucocorticoids is best for patients with NLR above 6 and the effect is largest in the region [6, 10].

Estimating heterogeneous treatment effects for all NLR values greatly reduces statistical power and inflates the width of the CIs. To test for statistical significance of the glucocorticoid benefit, we group NLR values into 3 ranges: NLR < 6, NLR 6-25, NLR >25. Using these three ranges, we estimate mean ARRs for NLR 0-6: 0.99(0.68-1.31), **NLR 6-25: 0.74(0.49-0.99)** and NLR > 25: 0.90(0.58-1.22). These values indicate that the benefit of glucocorticoids is statistically significant at p<0.05 for NLR 6-25, but not for either NLR < 6 or NLR>25.

Finally, while elevated NLR is a strong predictor of mortality, it is not the only risk factor (Figure S4). Our results suggest that glucocorticoids may have limited benefit to patients who are at high risk without having an elevated NLR. We encourage further investigation into treatments which could be effective for this set of patients.

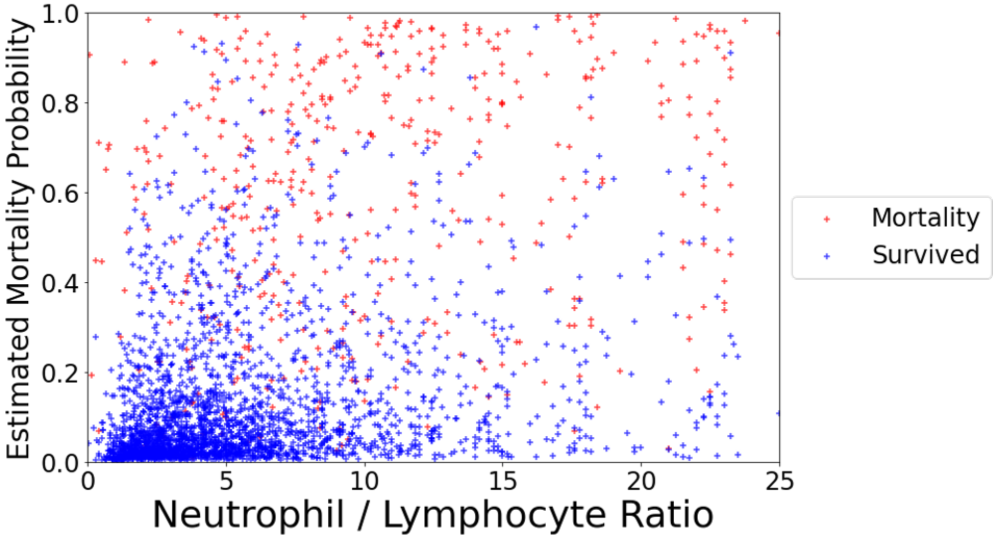


Figure S4: Estimated probability of mortality for each patient, as predicted by the risk model operating on patient features at admission. While there is a relation between elevated NLR and increased risk of mortality, there are a sizable number of patients who have a large probability of mortality without elevated NLR.

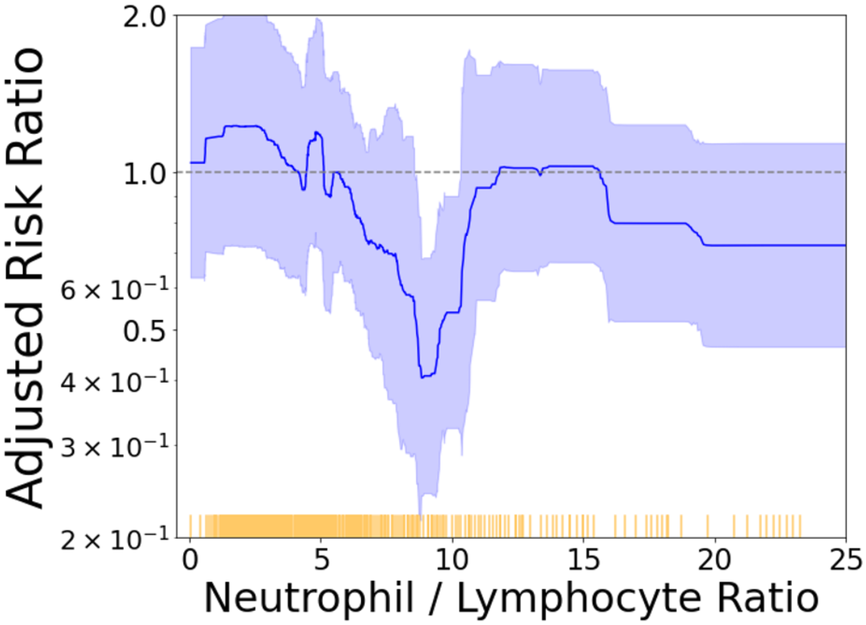


Figure S5: Estimated benefit (ARR) of glucocorticoid treatment after correcting for patient risk, stratified by NLR value. Shaded regions indicate 95% CIs. Despite the small sample size in each NLR bin, there is still statistical significance near NLR=8.5.

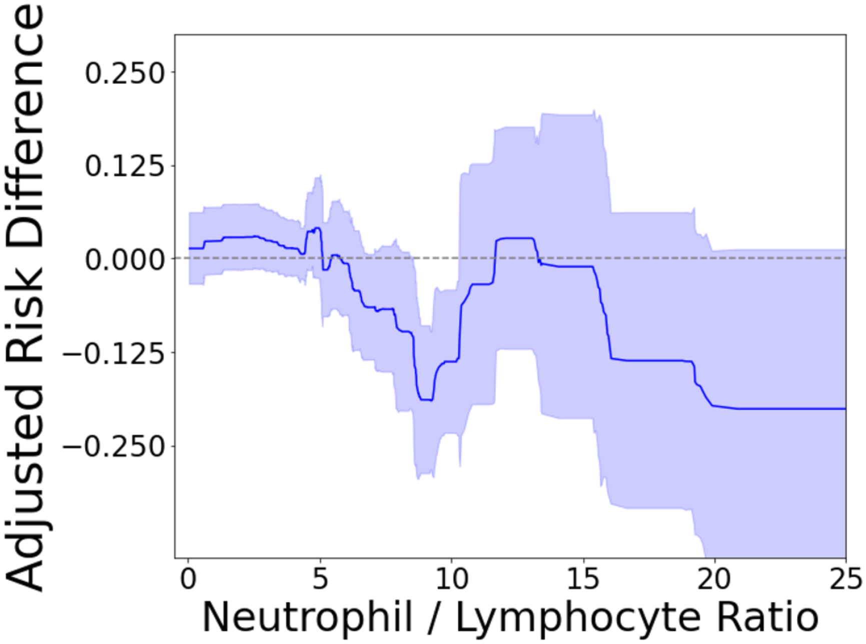


Figure S6: Estimated benefit (ARD) of glucocorticoid treatment after correcting for patient risk, stratified by NLR value. Shaded regions indicate 95% CIs. Despite the small sample size in each NLR bin, there is still statistical significance near NLR=8.5.
